# Inflammatory Type Focal Cerebral Arteriopathy of the Posterior Circulation in Children: a comparative cohort study

**DOI:** 10.1101/2023.04.15.23288630

**Authors:** Nedelina Slavova, Robin Münger, Iciar Sanchez-Albisua, Maria Regényi, Gabriela Oesch, Joël Fluss, Annette Hackenberg, Sébastien Lebon, Oliver Maier, Alexandre Datta, Sandra Bigi, Sebastian Grunt, Maja Steinlin

## Abstract

**Background:** Inflammatory type Focal Cerebral Arteriopathy (FCA-i) in the anterior circulation (AC) is well characterized and the FCA severity score (FCASS) reflects the severity of disease. We identified cases of FCA-i in the posterior circulation (PC) and adapted the FCASS to describe these cases.

**Methods:** Patients from the Swiss NeuroPaediatric Stroke Registry (SNPSR) with ischemic stroke in the PC and AC due to FCA-i and available neuroimaging were gathered. Comparison of data regarding Pediatric National Institutes of Health Stroke Scale (pedNIHSS) score and Pediatric Stroke Outcome Measure (PSOM) and FCASS was performed. We estimated infarct size by the modified pediatric Alberta Stroke Program Early Computed Tomography Score (pedASPECTS) in children with AC stroke and the adapted Bernese posterior diffusion-weighted imaging (DWI) score in the PC.

**Results:** Thirty-six children with a median age of 6.3 years ([IQR 2.8,8.6; range 0.9,15.6], 21 males, 58.3%) with FCA-i were identified. The total incidence rate was 0.151/100 000/year (95%CI 0.109–0.209). Seven had PC FCA-i and 5 had FCA-i in both circulations. Time to final FCASS was longer in the PC compared to AC, evolution of FCASS did not differ. Initial pedNIHSS was highest in children with FCA-i in the PC with a median of 8.0 (IQR 5.0-18.0), compared to 4.5 (IQR 2.0-8.0) in those with AC FCA-i and 6.0 (IQR 6.0-6.0) with involvement of both AC and PC. Different to the anterior cases PC infarct volume did not correlate with higher discharge, maximum or final FCASS scores (R 0.25, 0.35, 0.54).

**Conclusion:** The PC is affected in up to one third of cases of FCA-i. These cases should be included in future investigations on FCA-i. Although it did not correlate with clinical outcome in our cohort, the modified FCASS may well serve as a marker for the evolution of the arteriopathy in posterior FCA-i.

## Background

Focal cerebral arteriopathy of the inflammatory type (FCA-i) is an important risk factor for arterial ischemic stroke (AIS) in previously healthy children.^1^ After the first description as transient cerebral arteriopathy in 1998^2^, FCA-I was defined in 2004^2^ by a typical vascular imaging within 3 months after stroke showing unilateral focal or segmental stenosis or occlusion at a typical location, non-progressing on follow-up imaging 6 months post-stroke.^3^ Typical locations include the distal part of the internal carotid artery and the initial segments of the anterior, middle and posterior cerebral arteries (A1, M1 and P1 segments). As a special variant, post-varicella arteriopathy (PVA) was defined as TCA with a history of acute varicella infection within the 12 months before AIS.^4, 5^ Since then, herpes simplex virus, enterovirus, Mycoplasma pneumoniae or Borrelia burgdorferi infections have been reported as risk factors for FCA-i and AIS. ^6–11^

The term focal cerebral arteriopathy (FCA), which included unifocal or multifocal, unilateral or bilateral stenosis of variable etiologies (e.g. dissection or moyamoya disease and syndrome) was later defined, and TCA was seen as a subgroup of FCA^12, 13^. In the large international Vascular Effects of Infection in Pediatric Stroke (VIPS) study, the definition of FCA was narrowed to unifocal and unilateral stenosis or irregularities of the AC. The VIPS study further subdivided FCA into three groups: intracranial arterial dissection (FCA-d), inflammation (FCA-i) or undefined etiology (FCA-u)^1, 14^. FCA-i accounted for the majority of FCA in the VIPS study.^1^ The evolution of FCA-i is characterized by a monophasic natural course, frequently showing early progression (over days to weeks), which reaches a plateau with non-progression by 6 months. The majority of individuals improve or remain stable, but complete resolution is seen in only a minority of cases.^2, 3^

Based on the hypothesis of an inflammatory pathogenesis, corticosteroids are increasingly used to treat FCA-i.^15^ However, there is still no clinical trial data to support this treatment. The planning of forthcoming clinical trials in this field (Brechbühl et. al., PASTA Trial, unpublished data, 2023) revealed the need for a quantitative measurement of FCA severity. This led to the development of the FCA Severity Score (FCASS) based on a sub cohort of children with FCA enrolled in the VIPS study.^14, 16^ Only children with FCA in AC were included precluding its utility for posterior circulation stroke.

A few case reports have described PVA affecting the PC.^17–19^ A study on post-varicella AIS in Denmark included cases fulfilling clinical and radiologic criteria as well as a positive cerebrospinal fluid test for varicella-zoster virus (VZV) DNA in combination with intrathecal VZV IgG production. This includes one case of post-varicella AIS affecting the posterior circulation. ^20^ A population-based Swiss study revealed an incidence of PC AIS of 0.18/100 000 children /year. In 25.6% of these cases, the etiology was identified as FCA (FCA-i, FCA-d and FCA-u).^21^ There is currently no scoring system available to describe these cases in detail.

Children with FCA-i mostly present with hemiparesis or hemiplegia.^20, 22–24^ In general, children with AIS in the PC show less specific clinical symptoms and signs than those with AIS in the AC.^21^

The aim of our study was to identify all cases of FCA-i in the Swiss NeuroPaediatric Stroke Registry (SNPSR), to adapt the FCASS to encompass PC FCA-i and to compare clinical and radiological features of FCA-i between PC and AC.

## Methods

### Patients

We did a retrospective analysis of the prospective nationwide SNPSR which was initiated in January 2000.^22^ It comprises clinical and radiological data on every reported case of childhood AIS across Switzerland. Hospital-based pediatric neurologists are regularly reminded to report all pediatric stroke cases to the registry in Bern. The follow-up of these patients is organized centrally from Bern by questionnaires sent to the parents at 2 and 5 years and the clinicians are asked to send in reports from clinical follow up visits according to their local schedule. Short term follow up is usually done by a pediatric neurologist.

The SNPSR has been approved by the competent Ethics Committees and the Swiss Federal Ministry of Health. All registered patients or their legal guardians have provided written informed consent. Details of the SNPSR can be found in previous publications.^22, 25^ Some patients from the cohort have also been included in previous studies based on the SNPSR.^11, 21^

Childhood AIS was defined as a sudden (focal) neurological deficit with radiological proof of an acute ischemic lesion in the corresponding vascular territory. FCA-i included monophasic FCA, as in PVA and TCA.^26^

### Inclusion and Exclusion Criteria

All data of children (aged 1 month-16 years) with AIS registered between January 2000 until December 2018 with available good-quality neuroimaging at diagnosis and follow-up, with documented abnormalities on vascular neuroimaging, were retrospectively reviewed. In addition, charts from children with history of varicella infection within the last 12 months before the stroke were also reviewed.

Inclusion criteria were:

1. childhood stroke due to FCA-i defined by neuroimaging: focal stenosis, irregularity or banding in any vascular territory which was

a. either accompanied by vessel wall contrast enhancement on vessel wall imaging (VWI) or
b. preceded by infection (varicella within 12 months, other infections within weeks before stroke) and in the absence of trauma;
2. monophasic illness without progression on follow-up.

Exclusion criteria were arterial dissections, moyamoya arteriopathy, and underlying systemic disease like bacterial meningitis, neurofibromatosis, Kawasaki disease or systemic lupus erythematosus.

### Data collection

Demographic and clinical data, accompanying infectious risk factors and related laboratory investigations, as well as information on the patients’ treatment were searched for in the SNPSR. In cases of missing data, the original charts were reviewed. Data on the Pediatric National Institutes of Health Stroke Scale (pedNIHSS) score was collected as reported by the treating physician or performed retrospectively. Neuroimaging was retrospectively analyzed by a trained pediatric neuroradiologist (NS). Pediatric stroke outcome measure (PSOM)^27^ was collected from SNPSR or medical charts at discharge, after 6 months (range 3 to 9 months), and after 24 months (range 21 to 27 months). Outcome was dichotomized into good (total PSOM 0–0.5) or poor (total PSOM ≥ 1).

### Radiological Scoring of Arteriopathy Severity

FCASS was applied for assessment of the arteriopathy.^14, 16^ To measure severity of stenosis in the PC, the same principles were applied to the following segments: intracranial vertebral artery (VA), basilar artery (BA), P1 and P2 segments of the posterior cerebral artery (PCA), the superior cerebellar artery (SCA), the anterior inferior cerebellar artery (AICA) and the posterior inferior cerebellar artery (PICA), see STable 1.

FCASS was assessed on all MR angiography (MRA) and - if MRA was unavailable - on CT angiography (CTA). For further analysis, we included FCASS at baseline, maximum FCASS during follow-up (if more than one measurement available) and final FCASS (if follow-up imaging after > 6 months post stroke was available).

### Radiological Scoring of infarct Size

Infarct size on admission for AC stroke was calculated using the modified paediatric ASPECTS score^28–30^ (pedASPECTS), with a maximum of 30 points (15 for each side). Since pedASPECTS does not cover most locations in the PC, the modified Bernese diffusion-weighted imaging (DWI) score was used for stroke in PC,^31^ adding up to a maximum of 22 points (for details see STable 1).

### Statistical analyses

First, all variables gathered were analyzed descriptively. The incidence rate of FCA-i in PC in Swiss children was calculated using data published by the Swiss Federal Statistical Office^32^.

We compared the groups where the effects were strictly confined to the anterior or posterior circulation. Furthermore, we compared these cases to the group with mixed anterior and posterior involvement. If missing data could not be reconstructed retrospectively, patients were excluded from analysis. In-Between-group comparison was performed using nonparametric statistics (Wilcoxon rank sum and signed rank test). Binary outcomes were compared using Fisher’s exact test. Correlations between subgroups were evaluated with Spearman’s correlation coefficient. A 2-sided p-value < 0.05 was considered statistically significant. Analysis was performed using R statistical software, version 3.6.1.

We used the STROBE cohort checklist when writing our report^33^.

### Data availability

Anonymized raw data is available and can be shared upon reasonable request.

## Results

### Study Population and Epidemiology

During the study period from January 2000 to December 2018, 301 children with AIS aged between 1 month and 16 years were registered in the SNPSR (111 female [36.9%]; median age 5.8 years, IQR 1.9–11.4).

126 cases found in the registry with either documented vascular abnormalities on neuroimaging or an acute VZV infection within 12 months prior to stroke were reviewed clinically. In the first review all cases with a clear diagnosis other than FCA were excluded. The remaining cases underwent a central adjudication of the images.

Ninety children had to be excluded: In 9 cases, intracranial dissection was found to be the cause of FCA (FCA-d) and 11 cases were classified as FCA of undetermined etiology (FCA-u). In 10 cases extracranial dissection of the aorta or the cervical arteries was present, 1 case showed complete occlusion of the internal carotid artery of unknown etiology, and in 10 cases the arteriopathy was due to other types of pathology (e.g., systemic lupus erythematosus, Kawasaki syndrome, trisomy 21, neurofibromatosis type 1, moyamoya disease, and meningitis). Eight children had cardioembolic stroke and 9 children showed progressive central nervous system angiitis. In 32 cases no arteriopathy was found during expert radiological review.

FCA-i was found in 36 children. The total incidence rate of FCA-i in our Swiss cohort was 0.151/ 100 000 children/ year (95%CI 0.109–0.209). AC was affected in 24 children, PC in 7 with an incidence rate of 0.101 (95%CI 0.067-0.150), and 0.029 (95%CI 0.01-0.06) respectively. In 5 cases both circulations were affected with an incidence rate of 0.021 (95%CI 0.008-0.050).

Table 1 displays the baseline characteristics of all patients. Significant differences were found in the initial presentation: Patients with AIS affecting the AC presented more often with facial palsy than those with PC stroke. Headache and tetra paresis were significantly more often reported among cases with posterior FCA-i. Intravenous lysis was performed significantly more often if PC was affected. No between-group differences were found in inclusion criteria, demographic characteristics, and associations with previous infection or outcome.

**Table 1.**
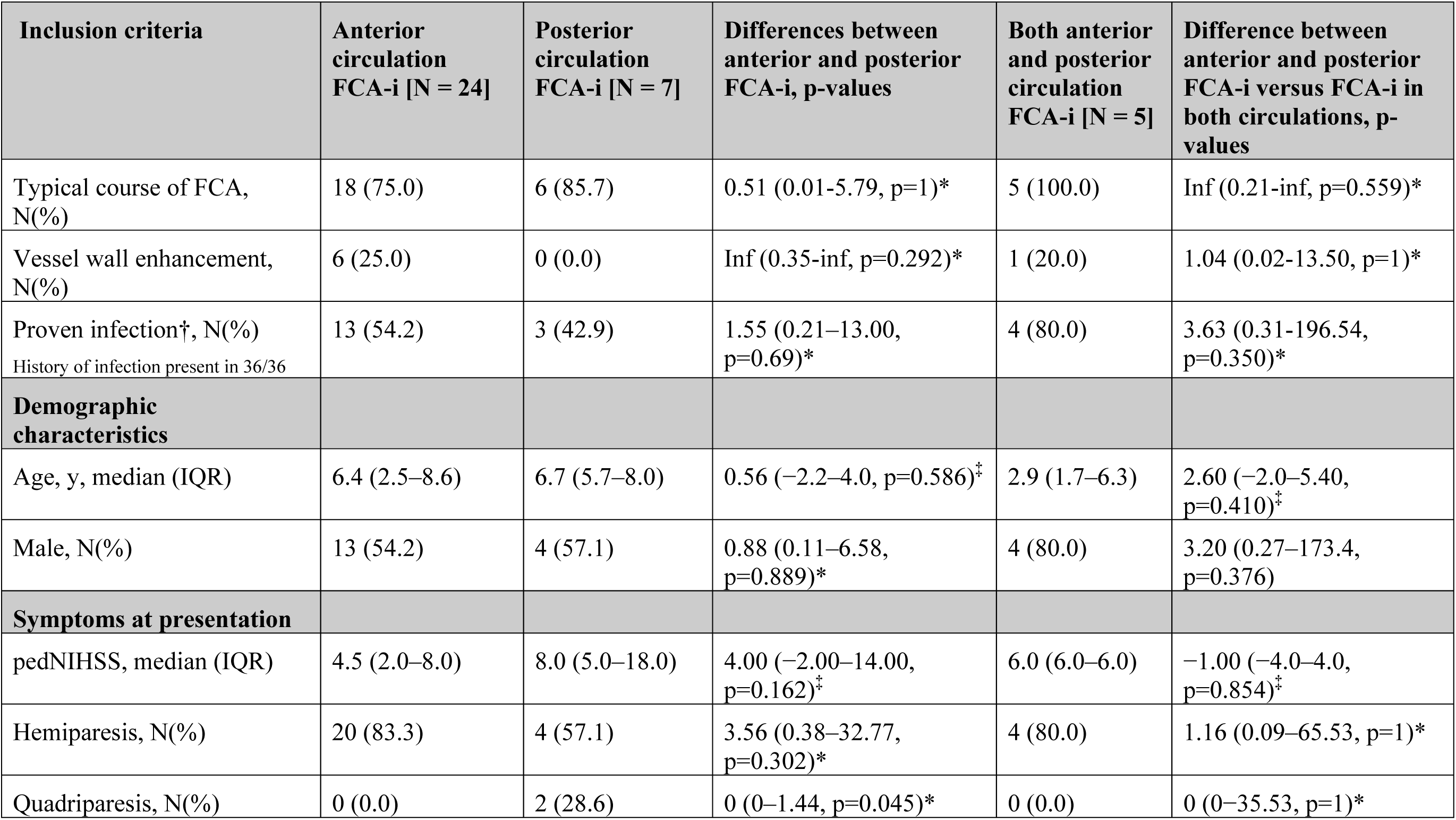
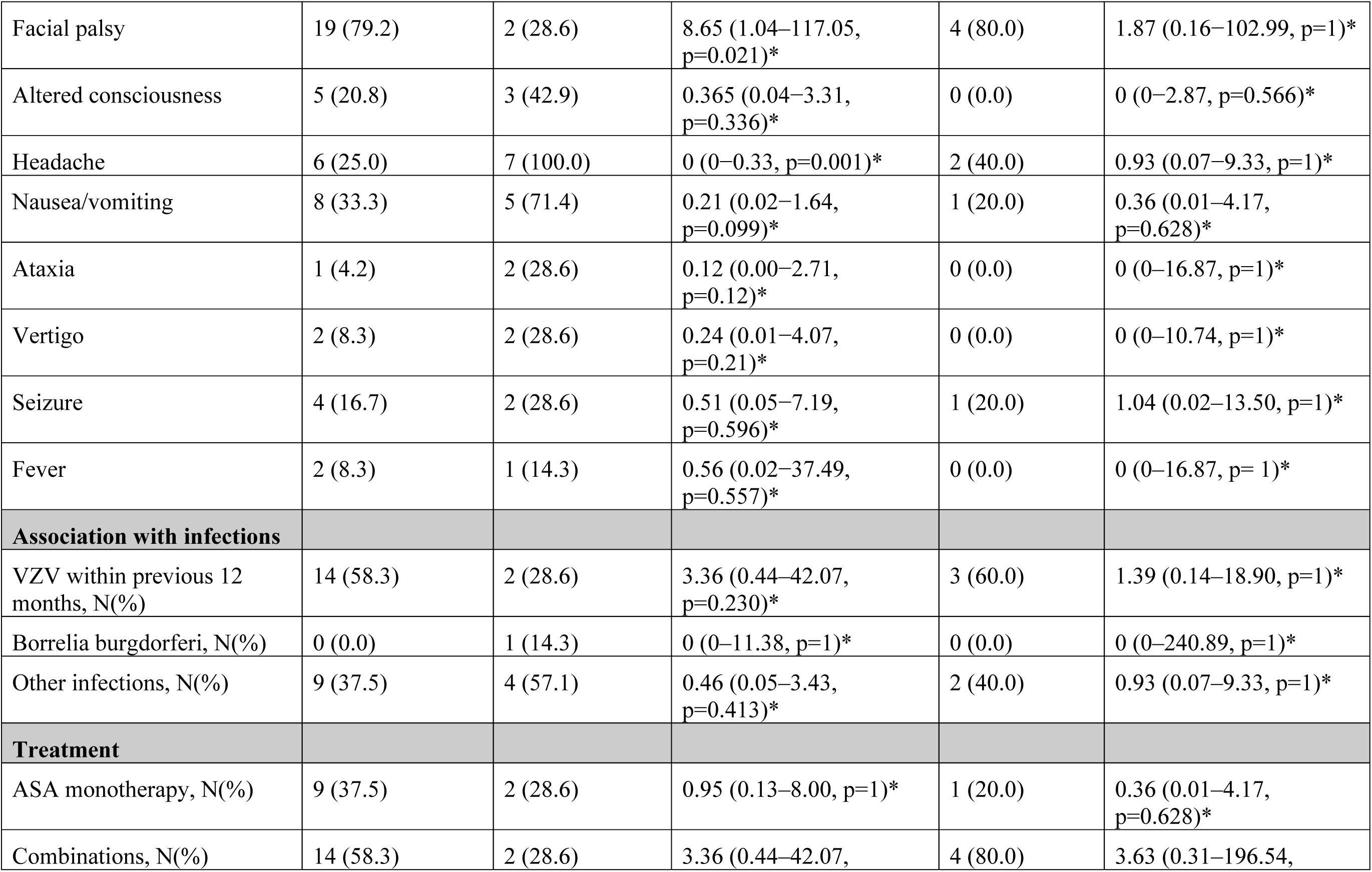
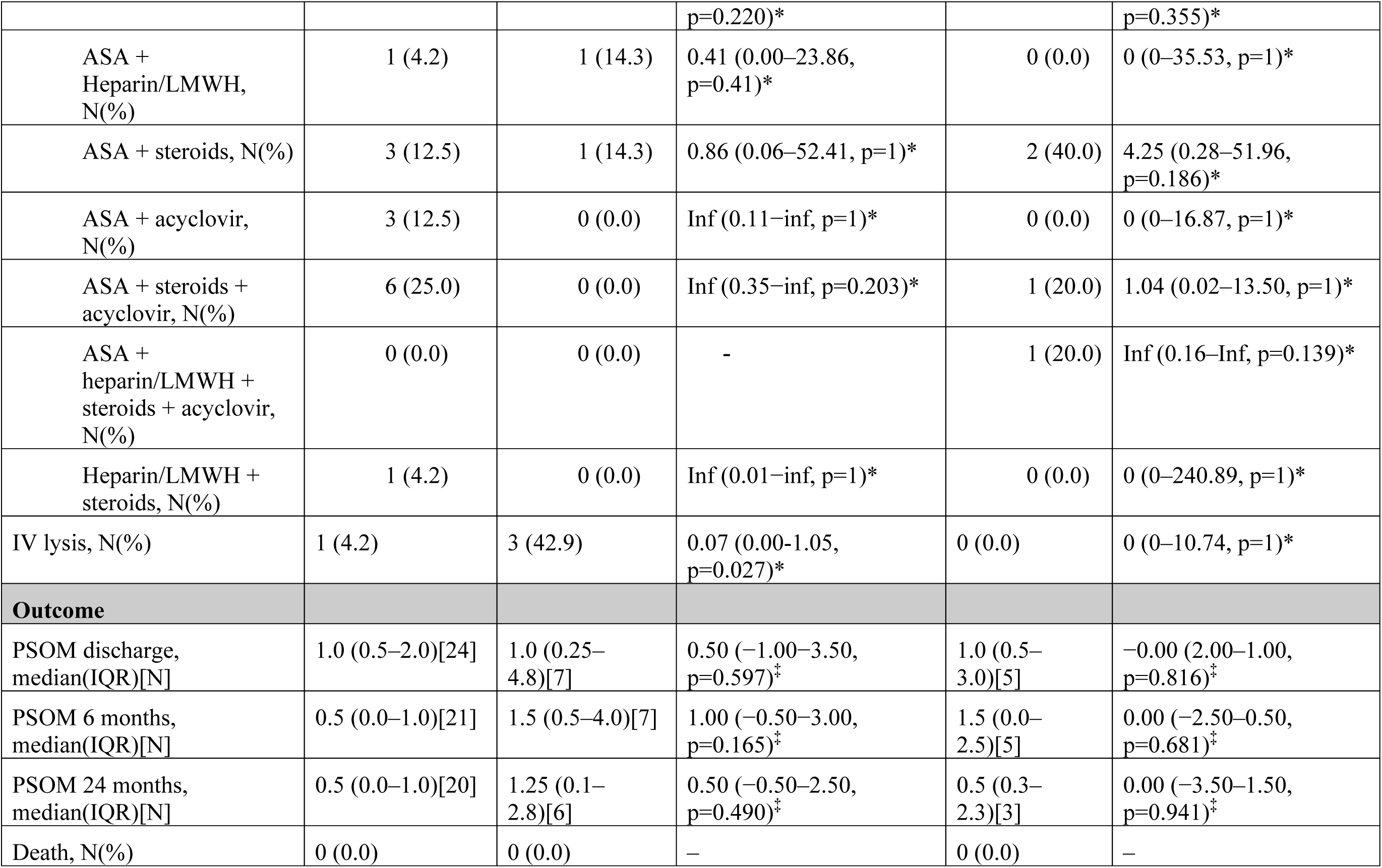
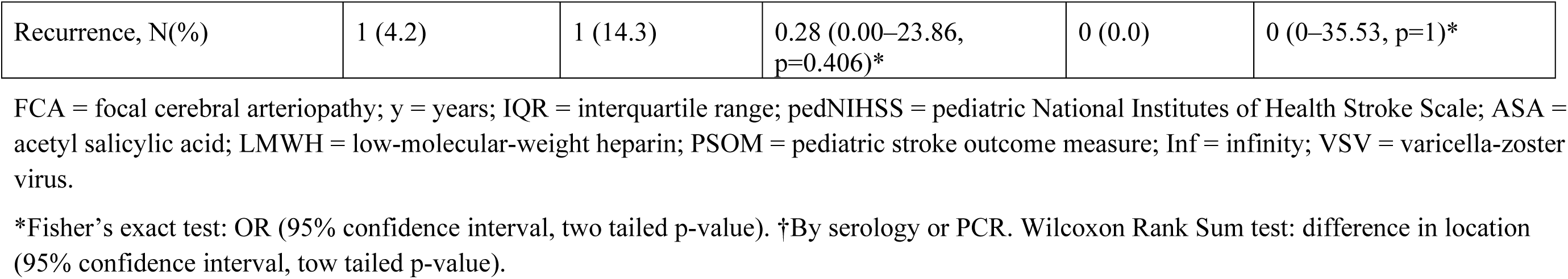
Summarized baseline characteristics of the patients

Children presenting with stroke affecting both circulations did not present with significant differences to the other two groups and this group was excluded from further comparisons.

Details on baseline characteristics, radiological findings, treatment, and outcome are summarized in STable 2.

### FCASS

Table 2 summarizes the FCASS obtained in our study for the anterior and the posterior group.

**Table 2.** Focal Cerebral Arteriopathy Severity Score (FCASS) summary at different time points

|  | Posterior circulation |  | Anterior circulation |  |
| --- | --- | --- | --- | --- |
|  | Time after stroke in days, median (IQR),[N] | Score, median (IQR) | Time after stroke in days, median (IQR),[N] | Score, median (IQR) |
| FCASS baseline | 1.0 (0.25–13.0),[6] | 9.5 (4.0–18.75) | 1.0 (0.0–10.0),[23] | 7.0 (3.5–12.0) |
| FCASS maximum | 15.5 (4.25–17.75),[6] | 12.0 (4.25–19.75) | 1.0 (1.0–7.5),[19] | 7.0 (5.5–12.0) |
| FCASS final | 2098 (1592–2458),[4] | 3.0 (3.0–5.0) | 451 (226–1185),[13] | 3.0 (0.0–5.5) |

FCASS baseline = FCASS of the first available imaging after stroke; FCASS maximum = highest FCASS of all available imaging – only for those patients with serial neuroimaging; FCASS final = FCASS of last imaging – only if imaging more than 24 weeks post-stroke was available; IQR = interquartile range; N = number of available FCASS in this group.

Baseline FCASS was obtained by MRA except in 3 cases of AC by conventional angiography and one case of PC by CTA (early years of the registry). All follow up FCASS were obtained by MRA.

As shown in Table 2, time to final FCASS differed significantly between the posterior and the anterior group with a median time of 300 weeks in the former (IQR 227–351, range 157– 359) and 64 weeks (IQR 32 - 169, range 24 - 545) in the latter group (p = 0.043). There was no difference in the scores or in the other time periods between the two groups.

Figure 1 shows the evolution of FCASS in the anterior and posterior circulation.

**Figure 1:**
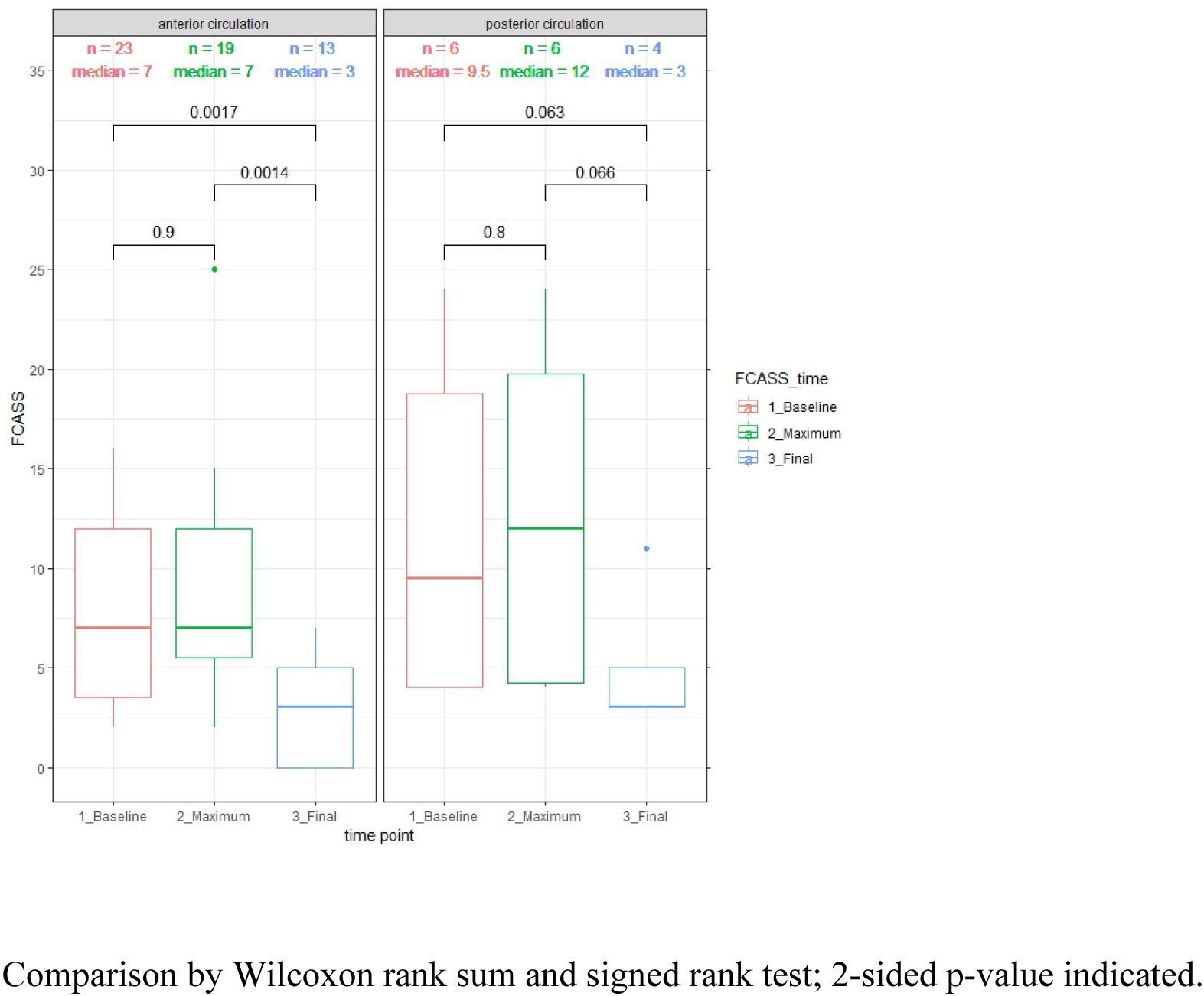
Baseline maximum and final FCAS-Score.

Improvement was evident after 6 months in all cases. The median improvement from baseline to final imaging in AC amounted to 4 points and 5.5 points in PC. Initial worsening seemed to be more pronounced if PC was affected, but there was no significant difference between groups.

Neither for the anterior nor the posterior group a correlation between pedNIHSS score and FCASS or PSOM and FCASS could be shown. There was also no significant difference between the groups in this regard.

No correlation was evident between FCASS and infarct size. Due to the different scoring system applied, direct comparison of infarct volume between the anterior and the posterior cases was not possible.

## Discussion

The purpose of this study was to characterize cases of inflammation-triggered FCA within the posterior circulation registered in the SNPSR and to compare them to cases of anterior FCA-i. Overall, PC was involved in one third of all FCA-i cases. In keeping with published data on PC-AIS in childhood^17–21^ we observed that FCA-i also affects PC in a non-negligible number of patients. Better understanding of the clinical presentation of these cases and application of a scoring system to assess severity of the arteriopathy is needed.^34^

Comparison of cases restricted to PC and AC showed a similar distribution of age. Sex distribution mirrored the typical pattern with more males affected by childhood AIS in general.^22^ Similar to PC stroke in general^21, 35, 36^ children with posterior FCA-i presented often with more non-specific symptoms, mainly headache. This group also showed a higher pedNIHSS at presentation. This might be because stroke in the brainstem will present with marked neurological symptoms. Facial palsy was found more often in the anterior group, consistent with the present literature.^21^

The association of VZV infection with FCA-i in the AC was not stronger than in PC. It is hypothesized that upon reactivation of VZV ganglia,^37^ the viral capsid might migrate through axonal retrograde transport (sensory nerve fibers) to the vessel wall of proximal segments of the circle of Willis, replicate and induce a local inflammatory response after transmural spreading. Experimental studies in monkeys demonstrated that intracerebral arteries receive afferent fibers from trigeminal and superior cervical ganglia (harboring latent varicella infection), not only to the carotid T-junction but to a lesser extent also to the posterior, rostral BA, PCA and SCA^38^ A study in rats demonstrated that the first and second spinal ganglion reach intracranial parts of the ipsilateral parts of the ipsilateral vertebral artery as well as the BA (as represented in Figure 2).^39^ This explains well the distribution of FCA-i in our study.

**Figure 2.**
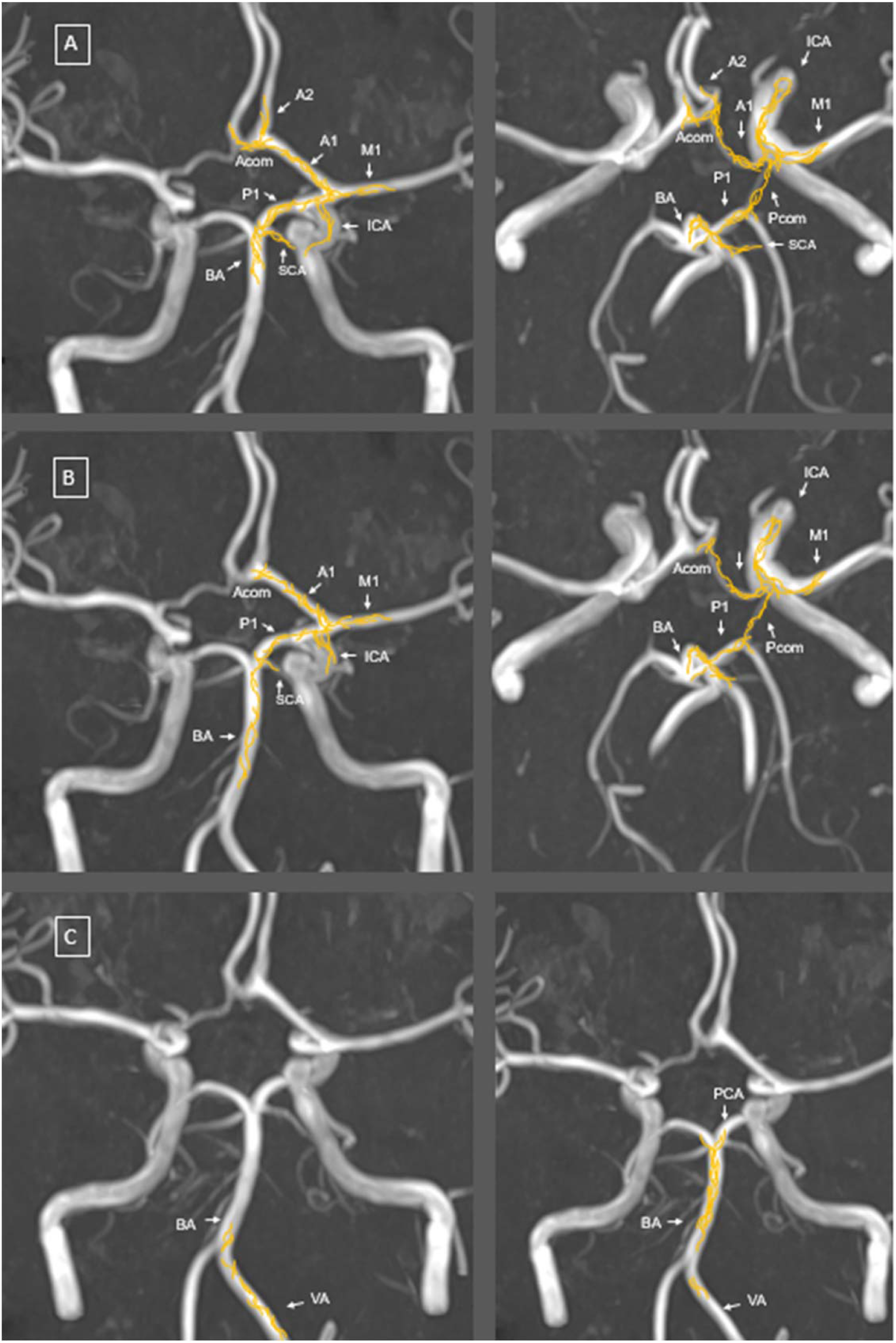
Schematic representation of nerve fibre distribution. Adapted from Arbab et al. using neuroanatomical tracing techniques after tracer injection into A) the trigeminal ganglion, B) the superior cervical ganglion and C) first (left image) and second (right image) spinal ganglia. The distribution of the anterogradely labelled nerve fibres (in yellow) was mapped on time-of-flight magnetic resonance angiography (MRA) maximum intensity projection (MIP) reconstruction imaging of the cerebral blood vessels. ACA, anterior cerebral artery; Acom, anterior communicating artery; BA, basilar artery; lCA, internal carotid artery; MCA, middle cerebral artery; PCA, posterior cerebral artery; Pcom, posterior communicating artery; SCA, superior cerebellar artery; VA, vertebral artery.

A major challenge is the distinction between FCA-d and FCA-i.^21^ In the early years of the registry FCA-i was not defined or scored. With the goal not to miss any case which in retrospect would be classified as FCA-i, initial inclusion criteria were taken very broad. This explains why so many cases had to be excluded upon clinical and radiological review.

All FCA cases were reviewed for the typical signs of dissections like ectasia, fusiform aneurysm, intimal flap, intramural hematoma or pseudoaneurysma. Consistent with the literature, history of head trauma was considered to support diagnosis of FCA-d.^1^ The presence of multiple lesions (as typical for PC cases) makes the diagnosis of a dissection very unlikely. Additionally, all lesions in our cases are localized in the typical region of the wandering of virus along sympathetic and sensory fibers as discussed before. In the cases affecting the BA the length of the lesion present is very atypical for an intracranial dissection. Considering all these points, we are convinced that our 7 cases in the posterior circulation are in fact FCA-i and with a high degree of confidence not FCA-d.

Concerning the difficulty of making a diagnosis of FCA-i we conclude that vessel wall imaging is needed if FCA-i is suspected, and special attention should be paid to the localization and the presence of multiple lesions.

Only one of our cases was associated with Lyme neuroborreliosis (LNB) and affected solely the PC, as most of the cases reported in the literature.^11^ Notably, this patient (case 25, STable 2) presented with mild symptoms only, but imaging studies revealed involvement of multiple arteries of the PC. This is in line with the pathophysiological hypothesis that the family Spirochaetaceae may induce perivascular inflammation rather than transport of infectious triggers via nerve fibers to vessels. A synopsis of published cases of LNB associated with pediatric stroke^11^ and our data strongly suggest that testing for LNB should be performed in cases of FCA-i-especially affecting the PC and multiple arterial segments.

Regarding treatment, intravenous thrombolysis was more often performed when the PC was affected, reflecting the severity of the acute disease, especially when BA was involved. Due to lack of randomized controlled trials on recanalization therapy in children with AIS, recommendations rely mainly on expert opinion.^40^ In the treatment of FCA-i, we consider that mechanical thrombectomy should be indicated very cautiously and only in selected patients with major vessel occlusion (e.g., carotid-T or M1). Interventions in vessels with underlying vessel wall inflammation may lead to unsuccessful recanalization and/or periprocedural complications. Intravenous or intraarterial thrombolysis (distant to local lesion) should be preferred over thrombectomy.

Median PSOM after 6 and 12 months was higher in the posterior group in our sample (Table 1), but the difference did not reach statistical significance, presumably due to the underpowered nature of the study. The median PSOM was 1 in cases of AC- and PC-FCA-i. This is in line with the original VIPS study.^41^

To score the extent of arteriopathy in the PC, we adapted the FCASS to fit PC-FCA. A validation of the posterior FCASS is not possible due to the small number of cases but would be desirable with a larger number of patients.

Previous FCASS studies have reported a positive correlation between infarct size and FCASS. The original FCASS study also found a correlation between higher 1-year PSOM and greater maximum FCASS, which could not be replicated in the Swiss study.^16^

We found no correlation between infarct size and FCASS or between PSOM and FCASS in cases involving the PC. Part of the explanation may be the different association between infarct size and clinical outcome in PC due to anatomical features and different dimensions. Further, the small sample size may well have prevented detection of possible correlations.

The most important limitation of our study is the small sample size, which makes comparison and correlation difficult. Even over this large time span, only a few cases of FCA-i within the PC were registered in Switzerland.

Because of the small sample size, significance levels must be interpreted with caution. However, we still consider our sample representative. Given the acquisition process and the well-established status of the SNPSR, we are convinced that most cases of childhood AIS during this time would have been captured. Thus, the conclusions drawn from our study still provide a solid basis for clinical decisions, highlighting the need for prospective randomized trials, especially regarding treatment regimens.

Another limitation is the lack of standardized guidelines for follow-up imaging. Since this is a retrospective analysis of a population-based registry and the patients were recruited over a long period of time, the availability of imaging studies and the imaging protocols have changed and differ between cases. To appreciate vessel wall enhancement, dedicated VWI is crucial, but was performed at presentation in only a minority of cases. According to the imaging recommendations, VWI is not necessarily performed in the acute setting^42^ although it can differentiate between the types of FCA.

We conclude that FCA-i occurs in the PC alone or in combination with FCA-i in the AC, although less often than isolated AC involvement. Thus, further studies in this field should consider including cases of FCA-i in PC. Children with posterior FCA-i present more often with non-specific symptoms. The modified FCASS may well serve as a marker for the evolution of disease but needs validation in another cohort. VWI is crucial in evaluating children presenting with PC stroke and should be included in stroke protocols.

## Data Availability

Anonymized raw data is available and can be shared upon reasonable request.

## Acknowledgements

We thank all participants of the Swiss NeuroPaediatric Stroke Registry as well as their parents.

We thank Heather J Fullerton (UCSF) for her valuable input in the discussion nd Mr. Charles Grose from University of Iowa Hospital, Iowa City to point our interest to the explanations of localizations of FCA-i; all the coworkers of the SNPSR: Andrea Capone Mori (Aarau), Alexandre Datta (Basel), Joël Fluss (Geneva), Annette Hackenberg (Zürich), Elmar Keller (Chur), Oliver Maier (St. Gallen), Jean-Pierre Marcoz (Sion), Claudia Poloni (Lausanne), Gian Paolo Ramelli (Bellinzona), Regula Schmid (Winterthur), and Thomas Schmitt-Mechelke (Lucerne).

## Supplemental Material

Supplemental Table S1a: Extended FCASS for the anterior and posterior circulation
Supplemental Table S1b. Modified Bernese Posterior Stroke Score
Supplemental Table S1c. Modified pedASPECTS
Supplemental STable 2: Detailed Results
Supplemental SFigure 1. Example of acute imaging in strictly posterior involvement

## Non-standard Abbreviations and Acronyms

AC: Anterior circulation
AICA: Anterior inferior cerebellar artery
AIS: Arterial ischemic Stroke
BA: Basilar artery
CI: Confidence intervall
CTA: Computed Tomography angiography
DNA: Desoxyribonucleic Acid
DWI: Diffusion-weighted imaging
FCA: Focal Cerebral Arteriopathy
FCA-d: Focal Cerebral Arteriopathy with intracranial arterial dissection
FCA-i: Inflammatory Focal Cerebral Arteriopathy
FCA-u: Focal Cerebral Arteriopathy undefined etiology
FCASS: Focal Cerebral Arteriopathy severity score
IQR: Interquartile range
MRA: Magnetic resonance angiography
PC: Posterior circulation
PCA: Posterior Cerebral artery
pedASPECTS: modified Pediatric Alberta Stroke Program Early Computed Tomography score
pedNIHSS: Paediatric National Institutes of Health Stroke Scale PICA Posterior inferior cerebellar artery
PSOM: Pediatric Stroke Outcome Measure
PVA: Post-varicella Arteriopathy
SCA: Superior cerebellar artery
SNPSR: Swiss NeuroPaediatric Stroke Registry
TCA: Transient Cerebral Arteriopathy
VA: Vertebral artery
VIPS: Vascular Effects of Infection in Pediatric Stroke
VWI: Vessel wall imaging
VZV: Varicella Zoster Virus

